# The lived experiences of diploma nurses who did not pass their professional licensing examination on the first attempt: A qualitative phenomenological study

**DOI:** 10.64898/2026.09.03.26362188

**Authors:** Kenneth Afful Adjei, Augustine Amoah, Richard Sakyi, Philip Abu, James Agamah Adabre, Daniel Cudjoe, Edward Appiah Boateng, Veronica Millicent Dzomeku

## Abstract

**Background:** Nurse educators in various health training institutions guide and prepare trainee nurses to be well equipped to write the Nursing Licensure Examination. They also draw on research in nursing education to shape their teaching practice. Most studies on nursing education to date have focused solely on predicting factors that may affect the academic performance of trainee nurses, with little to no attention given to the experiences that trainee nurses who are unable to pass their licensing examination encounter. This study presents the voices of trainee nurses who were unsuccessful at their first attempt at the licensing examination, their responses to failure, their perspectives of the factors that contributed to their failure, and the changes they made that led to subsequent success.

**Methodology:** A qualitative research methodology that employed a descriptive phenomenological approach was used to explore the lived experience of trainee nurses who were unable to pass their licensing examination on the first attempt.

**Results:** The results revealed common themes related to the psychosocial experiences, perceived causes of failure in the licensing examination, and successful remediation strategies for resitting the examination. The study revealed that shock, sadness, suicidal thoughts, social isolation, and a feeling of retardation are some of the psychosocial experiences that nurses who are unable to pass the licensing examination on first attempt encounter. Regarding the perceived causes of failure, participants acknowledged the role of personal behaviors such as prioritizing recreational activities over study time alongside distracting peer influences. To cope with and overcome failure, candidates relied on comprehensive family support (emotional and financial), encouragement from tutors and friends, spiritual support, and collaborative peer group studies using past examination questions, which ultimately enabled them to pass on their subsequent attempt.

**Recommendations:** The study recommends that health training institutions implement a program for nurses who are unsuccessful in the licensing examination to support their efforts to pass the examination on a subsequent attempt.

## Introduction

Global healthcare systems face a worsening human resource crisis driven by global demographic shifts, an aging population, and an escalating burden of complex, long-term chronic illnesses [1, 2]. Ironically, as the demand for qualified nursing care intensifies, the professional nursing workforce itself is rapidly aging, heightening the urgency to onboard highly skill nurses [3, 4]. The World Health Organization (WHO) projects a global deficit of 12.9 million nurses by 2035 [5]. This deficit is severely compounded by structural leaks in the educational pipeline, where a substantial proportion of graduate nurse trainees are unable to join the active clinical workforce due to their inability to pass professional licensure examinations [3, 6]. Because the nursing profession presents inherent structural risks to public safety if executed by undertrained personnel, standardized licensing examinations serve as the definitive, legally mandated gatekeeper ensuring that entry-level nurses are safe, skilled, and competent [6, 7].

While international benchmarks demonstrate high initial success such as in the United States, where the first-time pass rate for the NCLEX-RN averages 84.43% [8], the reality in developing healthcare systems is starkly different. In Ghana, the Health Professions Regulatory Bodies Act, 2013 (Act 857) explicitly mandates the Nursing and Midwifery Council (NMC) to secure the highest standards of training and practice in the public interest [9]. The NMC administers this high-stakes professional licensure examination biannually (in February and August). To transition into the workforce as Registered Nurses, diploma nurses (DNs) must conform to strict regulatory standards by passing three distinct papers: Medical Nursing, Surgical Nursing, and a comprehensive General Paper spanning Public Health, Pediatric, Psychiatric, Nursing Ethics and Obstetric Nursing. Under the current guidelines, candidates must achieve a minimum score of 50% on each individual paper; failing a single component invalidates the entire sitting, forcing candidates to re-register and incur full examination fees for subsequent attempts. Consequently, Ghana faces a critical clinical bottleneck: the national average first-attempt success rate hovers at just 50%, meaning half of all graduate DNs fail to transition into active clinical service annually [10, 11].

The consequences of initial licensure failure extend far beyond institutional performance metrics. In higher education, institutional success is traditionally measured by quantitative summaries like Grade Point Average (GPA) or mean rate evaluations [12, 13]. However, professional nursing education demands the simultaneous mastery of theoretical knowledge and clinical, hands-on skills [14]. Because nursing training institutions maintain highly competitive entry criteria, accepted candidates are widely presumed to be high achievers, a status that imposes immense societal and self-induced performance expectations [15]. When a candidate fails, the impact is multi-dimensional, fracturing the financial stability of parents and governments who funded the training, while simultaneously depriving healthcare facilities of critical bedside personnel [12, 16].

Extensive quantitative literature has historically investigated the systemic, external, and internal barriers to academic success within nursing education, categorizing them into student-related, home-related, institutional, clinical practice, and educator-related factors [14, 17]. Prior research indicates that poor learning attitudes, baseline testing anxiety, overwhelming domestic responsibilities, and weak institutional preparation are primary correlates of licensure failure [11, 16]. Yet, a critical gap in implementation and supportive science remains. While previous studies extensively explore these predictive variables, they overwhelmingly evaluate active student populations prior to graduation [14, 16, 17]. Once a diploma nurse graduates and fails the licensure examination, they exit the formal academic system. At this juncture, institutional support structures such as faculty tutoring, structured review coaching, peer study groups, and counseling services are drastically restricted or entirely absent [3, 10].

High-stakes examination failure is an deeply disruptive life event, and there is minimal comprehension regarding how post-graduate candidates phenomenologically process and make meaning of this failure during isolated remediation [15]. Initial licensure failure triggers severe psychosocial trauma characterized by social isolation, a profound loss of professional identity, compromised self-esteem, and acute guilt over disappointing family networks [4, 15, 16]. In highly competitive healthcare training environments, unsuccessful candidates are frequently stigmatized as weak individuals [15], putting them at elevated risk for psychological distress, depressive manifestations, shame, and suicidal ideation [10, 15].

How candidates cope with this distress directly dictates their long-term professional retention, subsequent test performance, and general well-being [11, 13]. Academic coping strategies typically bifurcate into adaptive problem-solving or maladaptive avoidance and mental disengagement [13]. While literature suggests interventions like targeted psychological counseling and collaborative partnerships between healthcare organizations and nursing programs [4], these strategies cannot be effectively deployed in sub-Saharan Africa without first mapping the specific, contextual realities of the affected individuals. Most existing qualitative studies have been carried out internationally, leaving a critical knowledge gap within the Ghanaian context. To design empathetic, effective systemic reforms, stakeholders including nurse educators, training college administrators, and health ministries must view licensure failure through the experiential lens of the candidates themselves.

Therefore, this study aimed to explore the lived experiences of diploma nurses in Ghana who did not pass their professional licensing examination on the first attempt. By providing an in-depth, phenomenological account of their psychosocial experiences, perceived failure factors, and subsequent coping mechanisms, these findings offer crucial insights to help stakeholders optimize remediation pathways and improve national licensure pass rates

## MATERIALS AND METHODS

### Ethical Consideration

All ethical protocols regarding human research were rigorously followed. Ethical approval was sought and obtained from the Committee on Human Research, Publications and Ethics (CHRPE) at the Kwame Nkrumah University of Science and Technology (KNUST) with reference number CHRPE/AP/179/20. Written informed consent was obtained from all individual study participants prior to data collection. Additionally, administrative permission was obtained from the management of the Nursing and Midwifery Training College, Sunyani, as well as the authorities of the specific hospitals where the participants were practicing. All foundational ethical guidelines concerning the use of human participants in research including voluntary participation, privacy, anonymity, confidentiality, and the principle of no harm were strictly adhered to throughout the research process. Data collected from respondents were kept under lock and key, accessible only to the researcher and supervisor, while soft copies were secured on a password-protected computer.

### Study Site

The study focused on diploma nurses who had been unable to pass their professional licensing examination on their first attempt at the Nursing and Midwifery Training College, Sunyani. At the time of the study, this cohort of nurses was completing their mandatory national service within two healthcare institutions inside the Sunyani Municipality of Ghana: The Bono Regional Hospital and the Seventh Day Adventist (SDA) Hospital. Both facilities address the health needs of the population within Sunyani and its surrounding environs, with the Bono Regional Hospital functioning as the primary referral center for all other health facilities within the Bono Region. While deployed at these facilities, the national service nurses performed clinical roles assigned by their superiors and provided routine feedback on their duties.

### Research Design

This study utilized a qualitative methodology with a descriptive, transcendental phenomenological approach to gain an in-depth understanding of the lived experiences of the participants [18, 19]. Rather than utilizing hermeneutic approaches that interpret actions in context [20], transcendental phenomenology focuses on the pure description of human experience. Consequently, the researcher’s personal experiences were not used to interpret the research questions. Openness was maintained during active questioning, allowing the investigator to access the private feelings and subjective reflections of the individuals who lived the phenomenon.

#### Sampling and Sampled Participants

The study population comprised practicing diploma nurses (DNs) who completed their pre-licensure education at the Nursing and Midwifery Training College, Sunyani, in the year 2018. This specific population consisted of individuals who were unsuccessful on their first attempt at the licensing examination, lived with that failure, prepared for the examination a second time, and successfully passed on their subsequent attempt.

A non-probability, snowball sampling technique was used to select and interview participants. This technique was strategically selected to locate a targeted population with unique characteristics, which can otherwise be difficult to identify due to the social stigma associated with examination failure [21]. The researcher determined that data saturation was achieved when successive participants provided identical or highly similar responses to the core questions. In total, ten (*N* = 10) participants were interviewed.

### Data Collection Approach

The primary data collection tool was an investigator-developed semi-structured interview guide consisting of two main sections: participant demographic information and a targeted question guide tailored to the study’s purpose. Pilot interviews were conducted with four (*n* = 4) diploma nurses who graduated from the Berekum Nursing and Midwifery Training College to identify structural flaws in the guide. This pretesting process facilitated the addition of strategic probing questions and sharpened the interviewing skills of the researcher [22]; data from these pilot interviews were excluded from the final analysis.

The definitive data collection was conducted via face-to-face semi-structured interviews, which facilitated access to diverse viewpoints and detailed descriptive insights [23, 24, 25]. Because all participants held a formal Diploma in Nursing, the interviews were conducted in English. Probing questions were continuously utilized to elicit full responses and structural clarifications. Interviews lasted between 30 and 45 minutes and were conducted at convenient, quiet locations chosen by the participants: the Sunyani Town Library and the Bono Regional Hospital conference room. Each interview was audio-recorded and transcribed verbatim. Ambiguous transcript segments were resolved by re-contacting the respective participant for clarity.

### Data Analysis

Data were analyzed using thematic analysis to identify and interpret patterns of experiences across the data set [26]. The analytic process was guided by classical thematic verification frameworks [27] and initiated immediately after the first interview. Each audio recording was transcribed verbatim and cross-examined by reading the transcript while listening to the audio.

The researcher immersed themselves in the data through constant reading to establish initial familiarization. Conceptual notes regarding items of interest and latent patterns were taken to generate codes. These coded and collated data extracts were then clustered to form potential themes of broader significance. The themes were systematically reviewed, refined, given suitable descriptive names, and synthesized into a textual description of the participants’ experiences regarding initial failure and subsequent preparation.

### Rigor and Trustworthiness

To establish qualitative rigor, four procedural criteria were applied: credibility, transferability, dependability, and confirmability [28]. Credibility was ensured by adopting an exploratory- descriptive approach aligned with the study’s core objectives, purposively selecting participants with direct lived experience of the phenomenon, and maintaining a reflective field diary to log nonverbal communications. Additionally, member-checking was deployed, whereby transcripts and extracted claims were returned to the participants to confirm their veracity [29]. Indirectly linked with credibility, dependability was achieved by keeping a detailed, documented audit trail of all operational data logs, field steps, and research procedures under lock and key [28]. To maintain confirmability, the final interpretations were rooted strictly in the raw concepts shared by the informants [30]. This neutrality was supported by raw audio recordings, field observation notes, and supervisor reviews

## RESULTS

### Demographic Characteristics of the Participants

A total of ten (*N* = 10) participants were successfully recruited and interviewed, consisting of six males (60%) and four females (40%). Notably, recruitment faced substantial barriers due to the extreme social stigma associated with initial licensing examination failure; eight prospective candidates (both males and females) who were approached explicitly declined to participate in the study. All ten included participants possessed a tertiary formal education, held a Diploma in Nursing Certificate, and were fluent in English alongside at least one indigenous Ghanaian language; consequently, all semi-structured interviews were conducted entirely in English.

Regarding the specific parameters of their initial academic deficiency, four participants suffered referrals exclusively in Medical Nursing, three in Surgical Nursing, two in the General Paper, and one encountered simultaneous referrals in both Medical and Surgical Nursing. At the time of data collection, all ten participants had successfully cleared their respective referred papers on their second examination attempt and had been fully deployed into active clinical spaces to fulfill their mandatory national service rotation rotations.

### Organization of Themes

Thematic analysis of the verbatim transcripts yielded four (4) major themes and eleven (11) corresponding sub-themes mapping the trajectory of licensure failure, isolation, and eventual remediation.

**Table 1.** Themes and Sub-themes.

| Major Themes | Sub-themes |
| --- | --- |
| **1. Social Experiences after Failure** | • Social isolation<br>• Feeling of being left behind |
| **2. Psychological Experiences after Exam Failure** | • Shock<br>• Sadness<br>• Suicidal thoughts |
| **3. Perceptions on the Cause of Failure** | • Acknowledgment of "the role of the self" as part of the journey to failure<br>• Peer influence |
| **4. Coping with and Overcoming Failure** | • Family support<br>• Support from tutors and friends<br>• Spiritual support<br>• Engaging in group studies |

(Source: Field Data, 2022)

#### Theme 1: Social Experiences after Failure

Participants detailed profound shifts in their interpersonal dynamics following their referrals. The core narratives focused on an inability to interact within traditional social networks to conceal the acute shame associated with public academic failure. This social retreat was heavily reinforced by a perceived sense of developmental stagnation, as candidates watched their successful classmates transition immediately into active clinical employment while their own professional access remained legally blocked.

#### Social Isolation

Participants described varying methods of self-imposed withdrawal from both neighborhood environments and established peer networks to avoid distressing conversations regarding their unsuccessful examination performance. In multiple instances, existing supportive relationships were completely severed:

> *“I isolated myself for a while to avoid uncomfortable questions… I was living in a self-imposed exile in my own family home.”* (P4)

> *“I couldn’t go close to my classmates again. I stopped going to church and other social activities.”* (P6)

Conversely, select participants intentionally restricted their social circles exclusively to fellow unsuccessful candidates, identifying them as the only safe, empathetic peer group capable of understanding the situation:

> *“It was those who had referrals that became my friends because I was not comfortable linking with those who passed.”* (P3)

#### Feeling of Being Left Behind

Participants expressed that initial licensure failure resulted in a complete suspension of their broader life trajectories, including the disruption of marital timelines, delays in transitioning into active wage-earning roles, and a general inability to plan for the future:

> *“We had already planned for our wedding, hmm. I did not care about it again.”* (P7)

The visual comparison of watching their immediate peers advance into clinical service while they remained structurally stagnant created deep feelings of professional regression and wasted academic effort:

> *“Looking at my friends moving in the same pace and me seeing them leading me, in fact, I saw it to be a step back, a very big step back… I feel like I have wasted three years because some of those people who passed are working and they are having their rotation service and I am in the house.”* (P2)

> *“I could not associate very well with my colleagues as they had all passed and now thinking ahead, I could not also fit into the association with my juniors.”* (P8)

### Theme 2: Psychological Experiences after Exam Failure

The psychological toll of high-stakes failure was characterized by an immediate state of emotional destabilization that developed into long-term psychological distress. This baseline trauma manifested through deep sadness, shock, and acute existential crises.

#### Shock

The initial disclosure of results triggered acute disbelief. Participants struggled to reconcile their baseline academic performance expectations with regulatory failure:

> *“So when the vice principal told me that I couldn’t make it in medicine, in fact, I thought it was a joke or something… So at that instant, to be frank between man and God, I was very shocked.”* (P2)

> *“I think I was among the first ten students in the country to check the results but to my surprise, I had a referral.”* (P3)

For others, the shock prompted structural, unanswerable internal questioning regarding the operational causes of their failure:

> *“So I took my slip and left for home. A lot of questions came to mind. How? Why? What happened?”* (P7)

#### Sadness and Suicidal Thoughts

Following the initial shock, participants experienced persistent sorrow and depressive symptoms that repeatedly hindered their focus during early remediation efforts:

> *“Anytime I tried doing something and I remembered it, in fact, it pulled me… In fact, it was very heartbreaking.”* (P2)

> *“Hmmm, I was broken by the results… the depression was too much.”* (P4)

> *“I sat on the floor and cried. I cried a lot because all my study group members passed. I always locked myself up in the room crying all day, all night.”* (P7)

For one participant, the severity of the emotional suffering directly escalated into an active consideration of self-harm, which was mitigated only by perceived maternal obligations:

> *“One day I even thought of suicide but because of my mum, I have to be there to support her. Hmm.”* (P6)

### Theme 3: Perceptions on the Cause of Failure

Upon structured retrospective reflection, participants did not view their failure as entirely external; they carefully evaluated their own operational actions alongside negative external environmental influences.

### Acknowledgment of “the Role of the Self” as Part of the Journey to Failure

Participants readily identified internal behaviors, underlying academic vulnerabilities, or specific testing errors that directly contributed to their negative outcome. This included a documented history of academic struggle or a failure to prioritize high-stakes preparation over recreation:

> *“So any time that we have this kind of football activities, I won’t forgo football and turn back to my, my, my books. For that one, whether we are having licensing examination before time, whether you are a principal, I won’t just mind you.”* (P1)

> *“I knew I was not that good academically compared to most of my friends.”* (P5)

> *“My failure may be due to deviation of the questions.”* (P7)

#### Peer Influence

Participants also identified maladaptive peer attachments and interpersonal relationships as primary distractions that systematically eroded their dedicated study time:

> *“Friends have been one of the contributing factors that has led to the collapse of many students or many candidates in this world. I am number one and I have been, I have also been a victim.”* (P1)

> *“I had this boyfriend who took most of my time.”* (P6)

### Theme 4: Coping with and Overcoming Failure

To navigate the remediation period and achieve success on their second attempt, participants relied on a mixture of external support networks, spiritual practices, and collaborative peer learning.

#### Family Support

Immediate family networks served as the baseline source of emotional stabilization and financial relief, directly buffering the candidate against the immediate costs of re-examination fees and the psychological weight of failure:

> *“My biggest motivation, let me say, is my mum. She just gave me all the necessary internal, physical, mental, spiritual, everything support that I needed…”* (P1)

> *“My parents’ support was paramount… my family gave me all the support I needed.”* (P5)

> *“So from the family side, I think I had enough psychological support, financial support, and the spiritual support.”* (P2)

#### Support from Tutors and Friends

Encouragement from institutional faculty and role models provided structural reassurance, restoring the candidates’ self-efficacy and confidence during test preparation:

> *“So I said if the support is coming from the family, the vice principal, and my friends also encouraging me, it made me to start to believe that I can still make things happen.”* (P2)

> *“Yeah, support came from friends, tutors, and family especially my wife. She was very supportive not forgetting my role model that is my childhood friend who is a nurse. This support was emotional and financial.”* (P3)

> *“My tutors were supportive and they made sure we were very prepared for the exam.”* (P5)

#### Spiritual Support and Engaging in Group Studies

Religious interventions, prayers, and scriptural texts served as internal stabilizing mechanisms to manage testing anxiety and maintain emotional endurance:

> *“My pastor encouraged me to pray a lot, so I prayed, listened to music, and was with someone that could make me laugh a lot.”* (P7)

> *“I held myself very well through the motivation from the principal, some of my tutors, some friends, and my family per se and some motivational quotes from the Holy Quran and the Bible even as well.”* (P2)

> *“I took to the word of God and prayers and I was okay.”* (P6)

Finally, transitioning from isolated learning into highly structured peer group settings allowed participants to resolve complex clinical concepts and access vital testing resources:

> *“The group studies gave me a wider scope of understanding that made me to solve some questions.”* (P2)

> *“I moved back to Sunyani from Bolga to study with friends.”* (P3)

> *“I had some past questions from my study group members that assisted me a lot…”* (P8)

## DISCUSSION

The primary objective of this study was to explore the qualitative lived experiences of diploma nurses (DNs) who were unsuccessful on their first attempt at the professional licensing examination in Ghana. The findings illuminate how academic failure in a high-stakes professional environment cascades into profound social isolation, psychological distress, and delayed life milestones, while simultaneously highlighting the vital roles that familial, institutional, and spiritual support networks play in fostering academic resilience and eventual remediation. Out of the ten participants who took part in this research, four were female, while several other female candidates initially approached explicitly declined to participate. This hesitation is likely attributable to the profound social stigma and professional marginalization associated with academic failure in healthcare fields, a gendered vulnerability well-documented in existing qualitative literature [31]. The age of the participants ranged from 23 to 27 years, with a mean age of 25 years.

Prior empirical research indicates that demographic factors including age, gender, level of enrollment, and baseline fear of failure exert a significant combined influence on undergraduate health science outcomes [13]. Within professional nursing education, age and maturity at the time of entry into a clinical program remain vital metrics; evidence suggests that nursing trainees who are older than 23 years upon admission demonstrate higher ultimate licensure pass rates [32]. This baseline trend is reflected inversely in the current cohort. With a mean age of 25 years at the time of the study, participants averaged approximately 22 years of age when entering their diploma programs. This pattern suggests that entering a demanding, high-acuity nursing curriculum below a maturity threshold may increase a candidate’s vulnerability to initial licensure failure. This dynamic is closely linked to psychological and cognitive maturity, as ultimate academic success depends heavily on a learner’s capacity to navigate adverse, high-stakes functional stress environments [33].

Beyond individual demographics, the structural disruption of personal milestones emerged as a distinct consequence of regulatory examination failure. This study found that unsuccessful candidates routinely deferred long-term marriage plans, and those with prior engagements actively paused these plans until professional licensure was achieved on their subsequent attempt. This finding closely parallels qualitative evidence where prospective partners or families required graduate nurses to clear their licensing examinations as a structural prerequisite to moving forward with marital commitments [3], highlighting how institutional failure stalls personal, non-academic life trajectories. Furthermore, the demographic composition of the sample showed that the majority (seven participants) were Akans and identified as Christians. This distribution directly reflects the geographic and cultural setting of the study in the Bono Region of Ghana, which is predominantly Akan-dominated, and aligns with broader national census metrics indicating that the majority of the Ghanaian population practices Christianity [34].

When evaluating the social experiences following this licensure failure, the dominant narratives shifted toward intense social isolation and a pervasive feeling of stagnation or “retardation” in life progression. Participants detailed the acute burden of trying to navigate a local society that expected them to be actively engaged in their mandatory national service, a highly visible milestone they could not fulfill due to their unsuccessful examination status. Consequently, candidates routinely hid from neighbors and families to avoid uncomfortable conversations regarding their referrals, resulting in the erosion or severing of established social relationships. In a foundational study on graduate nurses facing multiple NCLEX-RN failures, participants reported fractured relationships with family members, faculty, and their training programs, leaving them feeling completely abandoned [3]. Conversely, the current study revealed an important contextual contrast: while Ghanaian participants isolated themselves from peers who had successfully passed, their immediate families and nursing faculty remained a resilient source of functional support as they prepared to resit the exam. This divergence is likely explained by the duration and frequency of the failure; the cohort in the earlier study experienced chronic, multiple failures over an extended period [3], whereas participants in this study were navigating the immediate aftermath of a single, first-attempt failure, meaning they had not yet exhausted their familial and institutional support networks.

These localized support structures reinforce qualitative focus-group research emphasizing that establishing a strong, supportive relationship between faculty and students is a cornerstone for building academic resilience and high-quality remedial outcomes [35]. When new graduate nurses leave the structured environment of their training institutions, they often experience a crippling sense of isolation and actively look for external support to guide their licensure preparation [3]. Globally, professional licensure serves as the regulatory standard for governmental bodies to protect public health, safety, and consumer welfare. Because candidates are strictly barred from clinical practice until they pass the state licensing exam, unsuccessful trainees are forced into an involuntary period of stasis, watching their peers advance into professional roles. While educational institutions utilize multiple indicators to measure program efficacy, licensure pass rates remain the primary benchmark of institutional quality [32]. Confronting these social realities means candidates carry the psychological weight of their referral continuously [18, 24]. In this study, this dynamic manifested as an erosion of professional identity, a pervasive feeling of being left behind, a sharp decline in self-esteem, and voiced feelings of guilt driven by the fear of disappointing their families [4].

Though the emotional toll was heavy, human responses to missing critical life goals vary widely, and participants’ reactions appeared to undergo a strategic motivational shift over time [36]. Evidence indicates that emotional processing following failure can serve to increase determination and catalyze a desire to improve upon past performance [36]. While the psychological trauma initially induced shock, sadness, and vulnerability, the cognitive processing of these intense emotions during the remediation phase ultimately acted as a motivational driver for success on their second attempt [36]. In professional education, there is an institutional assumption that enrolled students will naturally maintain proficiency and complete their programs within standard timelines [37]. For many nursing trainees in Ghana, however, the state licensing examination represents a major structural bottleneck. Receiving news of a referral triggers immediate psychological shock and acute stress, shifting the candidate’s core baseline mood into profound sadness. This sequence supports established frameworks showing that individuals exist within continuous, fluid mood states that adapt to external trauma [38].

This profound sadness can carry long-term psychological risks, as clinical susceptibility is tied closely to experiences of acute loss and perceived failure in major life milestones [39]. In an assessment of undergraduates across higher education institutions, academic performance and the fear of exam failure were identified as primary predictors of severe stress and anxiety [40]. In the current study, this sadness manifested in diverse ways, ranging from emotional breakdowns during routine daily activities to total self-isolation, prolonged crying spells, and severe depressive episodes accompanied by active suicidal ideation. Because prolonged social isolation is heavily correlated with clinical depression [41], the occurrence of suicidal thoughts among candidates within this cohort is a critical finding. Higher education students exposed to severe, unbuffered stress are at an elevated risk for developing complex mental disorders and engaging in high-risk health behaviors, including self-harm [41]. The systemic implication is clear: without appropriate coping mechanisms and early psychological intervention, the compounding effects of social isolation, structural stagnation, shock, and prolonged sadness create an environment highly conducive to suicidal behaviors.

Regarding the perceived causes that led to this trauma, participants identified a complex mix of internalized self-blame and external peer influence. Many candidates expressed explicit self-blame upon reflecting on their preparation, noting that they fell victim to academic procrastination, spent excessive time on non-academic distractions (such as football matches), or mistakenly assumed they could compress their study timelines immediately before the exam. This reflects broader empirical evidence linking academic procrastination to systemic failure and poor behavioral outcomes [40]. Other participants acknowledged struggling with baseline academic performance while in school or cited a failure to properly interpret and respond to the specific formatting of the high-stakes exam questions. Concurrently, negative peer influence was identified as a primary behavioral driver of failure. Young adults rely heavily on peer networks for a sense of social belonging and validation, often conforming to group norms at the expense of academic responsibilities. Individuals who resist these non-academic group activities frequently face mocking or social exclusion from their peers [16]. These social pressures normalize distractive behaviors, eroding necessary study time and directly contributing to examination failure. This finding reinforces extensive research regarding the role of peer groups in driving behavioral conformity among emerging adults [42–46]. Participants expressed a strong retrospective belief that if they had insulated themselves from peer-driven distractions, they would have successfully cleared the examination on their first attempt.

To navigate the remediation period effectively, candidates turned toward protective external networks, citing family support as a cornerstone of their successful second attempt. While a “fear of failure” can negatively influence a student’s initial approach to studying [13], robust external validation can mitigate this anxiety. The vital role of family networks observed here aligns with empirical parent-empowerment frameworks, which demonstrate that parental competence, active involvement, and emotional support directly correlate with enhanced academic resilience and performance outcomes in children [47]. This interface became vital post-graduation, as structured academic environments naturally diminish upon leaving an institution, forcing the graduate nurse to operate as an independent learner [3]. This newfound autonomy can be highly challenging. To navigate this transition, participants in this study actively returned to their former institutional mentors and tutors for guidance. This behavior contrasts with previous findings where isolated candidates reported not knowing where to access remediation resources [3]. In Ghana, while enrolled trainees benefit from a highly structured curriculum, scheduled faculty interactions, and immediate feedback, these formal services disappear once a candidate fails and exits the program.

Faced with this institutional gap, seeking external spiritual counsel from religious leaders emerged as a vital parallel coping strategy for these independent candidates. Experiencing an examination referral can induce a psychological response similar to mourning a profound personal loss [48]. Consequently, the spiritual guidance, reassurance, and words of encouragement provided by religious leaders served to restore hope and rebuild the psychological resilience necessary for candidates to attempt the examination again. Finally, while some literature suggests that general approaches to study have only a limited capacity to predict precise academic performance grades [13], the targeted use of group study formats in this cohort proved highly effective. This finding contrasts with the wider conclusions of prior research [13], likely because those studies did not isolate the specific collaborative mechanics of peer-remediation groups. Emerging adults naturally seek group membership and show a high readiness to conform to the directions and expectations of valued peers within those groups [6]. When directed toward clear, shared academic goals, these structured study groups provide an exceptionally effective framework for achieving positive licensure outcomes.

## Conclusion

Licensing examination failure inflicts a profound, multi-dimensional burden on diploma nurses in Ghana, triggering a complex combination of emotional distress, social disruption, and personal setbacks. Unsuccessful candidates experience significant psychosocial strain, characterized by intense social isolation, a perceived stagnation in life progression, disrupted relationship and marital plans, and a damaged professional identity. The psychological toll is particularly severe, often manifesting as acute shock, profound sadness, depression, and critical mental health vulnerabilities, including suicidal ideation.

While participants attribute their failure to a combination of internalized self-blame regarding poor time management and negative peer influences that compromised their study focus, their path to recovery reveals a distinct dynamic. Overcoming this setback during subsequent attempts is heavily dependent on a multi-faceted support system. Ultimately, mitigating the impact of licensing failure requires comprehensive institutional interventions that look beyond traditional academic remediation. Nursing training institutions must implement structured counseling and formal support frameworks alongside family and spiritual networks to safeguard candidate well-being and facilitate successful entry into the healthcare workforce.

### Limitation of the Study

A limitation of this study is its specific temporal bounding, as data collection occurred exclusively in 2022. The lived experiences, emotional stressors, and systemic challenges reported by these trainee nurses reflect the specific educational, institutional, and regulatory environment of that post-pandemic period. Because this study captured a cross-sectional snapshot rather than a longitudinal trajectory, these findings may not account for subsequent structural reforms in nursing curriculum, changes to licensing examination formats, or evolving institutional support systems implemented after 2022. This notwithstanding, Lincoln and Guba [28] asserted that the findings of qualitative research should be measured in terms of its transferability.

## Data Availability

All relevant data are within the paper and its Supporting Information files.

## Acknowledgments

We thank the trainee nurses who gave us their time and the needed information for this work.

## Author contributions

**Conceptualization:** Kenneth Afful Adjei

**Data curation:** Kenneth Afful Adjei, Augustine Amoah, Richard Sakyi

**Formal analysis:** Kenneth Afful Adjei, Augustine Amoah, Richard Sakyi, Philip Abu, James Agamah Adabre, Daniel Cudjoe

**Investigation:** Kenneth Afful Adjei, Augustine Amoah, Philip Abu

**Methodology:** Kenneth Afful Adjei, Augustine Amoah, Richard Sakyi, Philip Abu, James Agamah Adabre, Daniel Cudjoe

**Project administration:** Kenneth Afful Adjei

**Resources:** Kenneth Afful Adjei

**Supervision:** Veronica Millicent Dzomeku, Edward Appiah Boateng

**Validation:** Kenneth Afful Adjei, Augustine Amoah, Richard Sakyi, Philip Abu, James Agamah Adabre, Daniel Cudjoe

**Visualization:** Kenneth Afful Adjei, Augustine Amoah, Richard Sakyi, Philip Abu, James Agamah Adabre, Daniel Cudjoe

**Writing – original draft:** Kenneth Afful Adjei, Augustine Amoah, Philip Abu.

**Writing – review & editing:** Kenneth Afful Adjei, Augustine Amoah, Richard Sakyi, Philip Abu, James Agamah Adabre, Daniel Cudjoe

